# Covid-19 vaccine perceptions in Senegal and in Mali: a mixed approach

**DOI:** 10.1101/2021.10.06.21264664

**Authors:** Eleonore Fournier-Tombs, Massamba Diouf, Abdine Maiga, Sylvain Faye, Tidiane Ndoye, Lalla Haidara, Moussa Batchily, Céline Castet-Renard

## Abstract

This paper presents the results of two qualitative surveys in Senegal and in Mali, which include questions about hesitancy to the COVID-19 vaccine between April and June 2021. It took place within a larger 2-year research project involving researchers in Senegal, Mali and Canada which examines the uses of artificial intelligence technologies in the fight against COVID-19. The study involved 1000 respondents in Senegal and 555 in Mali. The researchers found that overall, 55% of respondents in Senegal and 52% of respondents in Mali did not plan to be vaccinated. Hesitancy was much higher in youth aged 15-35 in both cases, with 70% of youth in Senegal and 57% of youth in Mali not planning to be vaccinated, compared to only 42% of elderly in Senegal and 37% of elderly in Mali. The researchers did not find disparities between male and female respondents in Senegal but found some in Mali. They also found that those who had a member of the family with chronic disease (diabetes or hypertension) were slightly more likely to want to be vaccinated. Reasons for vaccine hesitancy fell in several categories, including fear of vaccine side-effects, disbelief in vaccine efficacy or usefulness, and general distrust in the public health system.

## Vaccination progress in Senegal and in Mali

In 2021, several vaccines mostly stemming from the Covax initiative were approved in Senegal – the Astra Zeneca vaccine, which is produced by Oxford University in England, the Johnson & Johnson vaccine, produced by Johnson & Jonhson in the United States, and the BBIBP-CovV vaccine, which is produced by Sinopharm in China^1^. In Mali, the Astra Zeneca vaccine is also approved, as is the Sputnik V vaccine, produced by Gamaleya in Russia^2^. As of October 6, 2021, 5.6% of the population of Senegal was fully vaccinated, with 1.8 million individual doses given, for a population of 16.3 million. In Mali, 1.1% of the population was fully vaccinated, with 413,000 individual doses given, for a population of 19.66 million in the same period (Reuters, 2021).

Low vaccination rates in these countries, however, are largely not due to vaccine hesitancy, but rather to vaccine access. The World Health Organisation (WHO) has raised concerns about lack of access to the vaccines in developing countries, noting in July 2021 that this issue was the largest barrier to the recovery from the COVID-19 pandemic (WHO, 2021). According to the UNDP’s Covid-19 data dashboard, 51.75% of people in high income countries and 1.36% of people in low-income countries had been vaccinated by August 9, 2021 (UNDP, 2021). Senegal and Mali have low vaccination rates even when compared to other countries in the low-income grouping.

## Vaccine hesitancy in Senegal and beyond

In a survey conducted between December 2020 and January 2021, Ridde et al (2021) found that a weak majority – 54.4% - of Senegalese respondents were planning to be vaccinated. A study by Johns Hopkins (2021) in Nigeria found that vaccine acceptability had increased slightly over time – from 54.7% at the beginning of the pandemic, to 61.3% in March 2021. A study by the research group CORAF listed the following reasons for vaccine hesitancy in Senegal: (1) safety and secondary effects; (2) the vaccine as a part of a conspiracy for population control; (3) the vaccine as an effort to profit financially from the pandemic; (4) preference for other kinds of protection, such as traditional medicine, alternative medicine or religion; (5) refusal of all vaccines; and (6) the vaccine as a method to collect data or conduct research about Africans.

Thiongane (2021) argues that vaccine hesitancy in Sub-Saharan Africa is nothing new in a context where there have been failures on the part of public health. In Niger, for example, a meningitis epidemic between 2015 and 2017 led to school closures and a vaccination campaign. Lack of sufficient vaccines caused enormous line-ups at pharmacies, where the price of the vaccine soon tripled. Counterfeit vaccines then appeared on the market, leading eventually to a parliamentary inquiry.

However, lack of trust in public health authorities as a reason for vaccine hesitancy is not unique to Sub-Saharan Africa. In a 2018 study on 1,173 patients in France, Meredith and Sivry (2018) found that 63% of respondents had some form of hesitancy in relation to vaccines, even though 90% agreed that vaccines could protect their children against severe illnesses. The reasons cited then echoed those found in this study, with 46% of the hesitant answering: “the vaccine is not safe”, and 21.8% noting: “the vaccine is not useful”.

While vaccine hesitancy was already a public health issue before the pandemic (MacDonald, 2015), it has become a global issue, namely because global recovery from COVID-19 has been tied to successful vaccination programs. Solís et al (2021), in a study of vaccine hesitancy in low- and middle-income countries, found high rates of vaccine acceptance. However, concern about side effects was the most common reason for hesitancy by respondents. Another study by the Africa Centres for Disease Control (2021), which included Senegal but not Mali, found that 79% of respondents in the countries surveyed would be vaccinated against COVID-19 if it was proven safe and effective.

This study therefore provides additional insight into the attitudes towards the COVID-19 vaccine in Senegal and in Mali, which have been the subject of few such studies, while still having largely unvaccinated populations.

## Survey description

The surveys were conducted in April 2021 in Senegal and June 2021 in Mali, by a group of local researchers trained by investigators from Université Cheikh Anta Diop (UCAD) in Dakar and CERCAD research centre in Bamako. In Senegal, 1000 individuals were surveyed, aged 18 and older, selected according to a three-stage sampling method of regions, households and individuals in six regions. The regions were chosen according to the higher number of COVID-19 cases compared to other parts of the countries, and included Dakar, Thiès, Diourbel, Saint-Louis, Ziguinchor and Kédougou. Each of the selected region had significant COVID-19 infection rates during at least one wave since March 2020. There was a strong urban representation in these samples, with 6 of the 10 largest cities in Senegal represented in this sample, including Dakar, with a population of approximately 2 million. These choices were guided by the evolutionary dynamics of COVID-19, which has had a more severe impact on urban areas.

In Mali, a similar approach and criteria was used for the selection of regions, households and individuals. Bamako and Koulikoro, both urban areas, have been heavily impacted by COVID-19. However, these were also selected due to concerns about the safety of the research team. 550 subjects aged 18 and older were surveyed in these areas.

The two sample sizes were calculated using the Schwartz formula, with allocation in proportion to the population of each region selected. The research mobilized nearly 50 investigators and interviewers in both countries, who were trained beforehand to have the same understanding of the questions and a uniform way of administering the collection tools to the people to be interviewed. Questionnaires and interview guides were used to collect information on socio-demographic characteristics and vaccination, among other questions.

**Figure 1:**
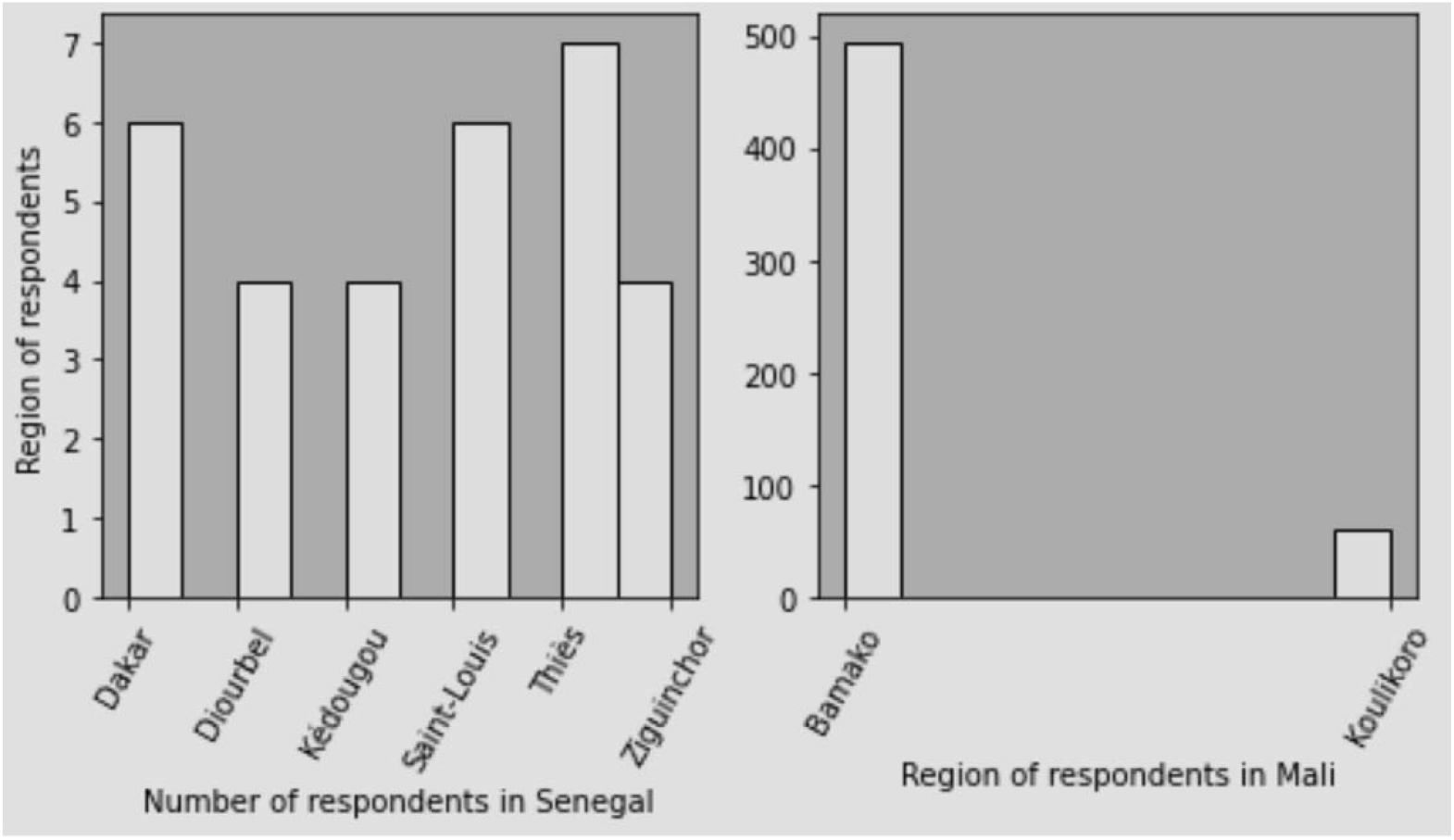
Survey respondent count by region in Senegal and in Mali

## Analytical methodology

This analysis relies on two analytical techniques: (1) descriptive statistics of respondent preferences and demographics; and (2) a topic analysis of their qualitative responses to question #3. The topic analysis methodology selected was Latent Dirichlet Allocation (LDA), a commonly used text analysis technique which groups texts according to the similarity of their contents, first described by Blei et al (2003).

To analyse the responses, we first created a database for all respondents who answered that they would either not get vaccinated or they were hesitant to do so. We then extracted the reasons for their hesitancy. In Senegal, there were 507 individual responses, and in Mali, there were 243. We then proceeded with the same analytical technique for both datasets.

In the pre-processing phase, we removed the stop words, which are a list of terms that were considered meaningless in the dataset, such as “I”, and “because”. We then created a sparse matrix of terms using term frequency-inverse document frequency (TF-IDF) which provided a value for each term based on its importance in the dataset, rather than a count value. Finally, we created an LDA model which would separate the responses into 6 categories, mimicking the number of response categories found in the CORAF (2021) study. The original analysis resulted in three distinct clusters, and a fourth grouping of the remaining clusters. We therefore ran the analysis again with 3 categories to improve the distinction between the clusters.

We also performed a similar analysis of terms used by those who were favourable to vaccination. The datasets were smaller – 367 individual responses for Senegal and 267 for Mali.

## Findings

The tables below show that 55% of those surveyed in Senegal and 52% of those surveyed in Mali considered themselves unlikely to get the vaccine. Of these groups, youth were more likely to display hesitancy than adults and the elderly, and women were less likely to display hesitancy than men.

**Table 1:**
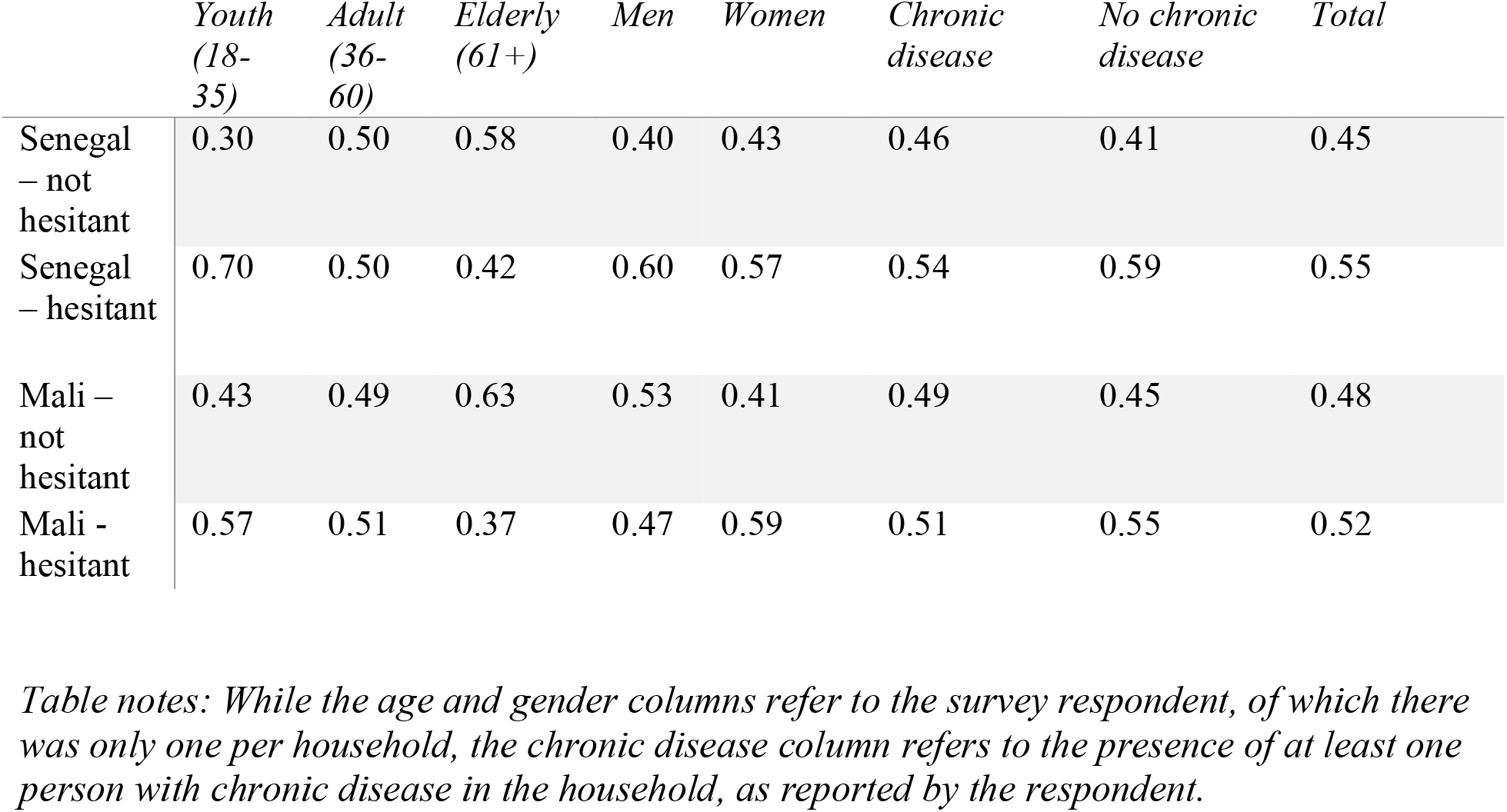
Vaccine perceptions by age, gender and chronic disease in Senegal and Mali

|  | *Youth*<br>*(18-35)* | *Adult*<br>*(36-60)* | *Elderly*<br>*(61+)* | *Men* | *Women* | *Chronic*<br>*disease* | *No chronic*<br>*disease* | *Total* |
| --- | --- | --- | --- | --- | --- | --- | --- | --- |
| Senegal – not hesitant | 0.30 | 0.50 | 0.58 | 0.40 | 0.43 | 0.46 | 0.41 | 0.45 |
| Senegal – hesitant | 0.70 | 0.50 | 0.42 | 0.60 | 0.57 | 0.54 | 0.59 | 0.55 |
| Mali – not hesitant | 0.43 | 0.49 | 0.63 | 0.53 | 0.41 | 0.49 | 0.45 | 0.48 |
| Mali - hesitant | 0.57 | 0.51 | 0.37 | 0.47 | 0.59 | 0.51 | 0.55 | 0.52 |

*Table notes: While the age and gender columns refer to the survey respondent, of which there was only one per household, the chronic disease column refers to the presence of at least one person with chronic disease in the household, as reported by the respondent.*

**Table 2:** Vaccine perceptions by level of instruction in Senegal and Mali

|  | *Instruction level 1, 6, 7* | *Instruction level 2* | *Instruction level 3* | *Instruction level 4* | *Instruction level 5* |
| --- | --- | --- | --- | --- | --- |
| Senegal – not hesitant | 0.33 | 0.43 | 0.44 | 0.40 | 0.49 |
| Senegal – hesitant | 0.67 | 0.57 | 0.56 | 0.60 | 0.51 |
| Mali – not hesitant | 0.43 | 0.45 | 0.47 | 0.47 | 0.46 |
| Mali - hesitant | 0.57 | 0.55 | 0.53 | 0.53 | 0.54 |

*Table notes: The instruction levels were categorized as follows in the survey: (1) No instruction; (2) Primary instruction (ages 6-11); (3) Secondary instruction (ages 12-17); (4) Superior instruction (college or university); (5) Koranic instruction; (6) Literacy only; (7) Other*

## Topic analysis

The reasons for vaccine hesitancy in both countries are presented here. In Senegal, the LDA topic analysis yielded three distinct topics, which are presented in the table below. The ten most frequent keywords for each topic are listed, as well as the percentage of responses that contributed to each topic. As we can see, there is overlap in topics, as certain respondents cited several different reasons for vaccine hesitancy.

**Table 1:** Reasons given for vaccine hesitancy in Senegal

| *Topic #* | *Top 10 keywords (translated from French)* | *Topic percent contribution* | *Representative response* |
| --- | --- | --- | --- |
| 1 | Fear, certainty, vaccine, rumors, uncertainty, good, hearsay, confidence, too much, wearing (mask) | 0.7214 | Because I am already healthy |
| 2 | Confidence, effects, secondary, again, lack, illness, causes, readiness, certainty, uncertainty | 0.7679 | It's all about politics |
| 3 | Reassuring, doubt, informed, efficacy, will, good, wait, confidence, inside, risks | 0.7484 | I prefer other protection measures |

**Table 2:**
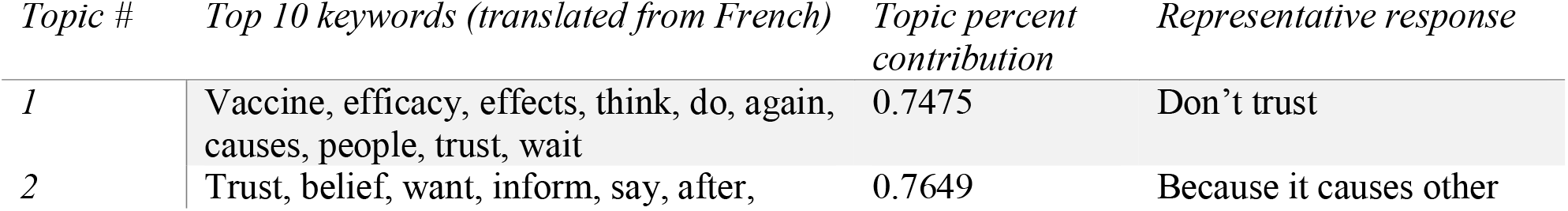
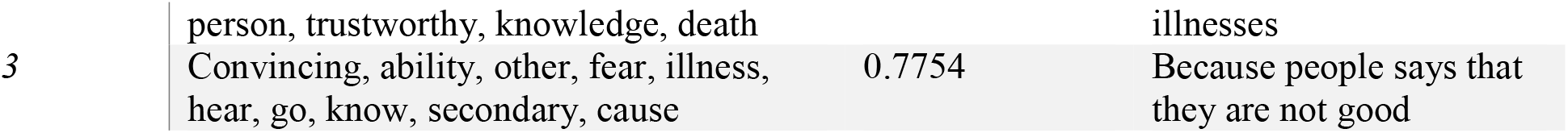
Reasons given for vaccine hesitancy in Mali

The tables below show the reasons behind vaccine acceptance in Senegal and in Mali.

**Table 3:** Reasons given for vaccine acceptance in Senegal

| *Topic #* | *Top 10 keywords (translated from French)* | *Topic percent contribution* | *Representative response* |
| --- | --- | --- | --- |
| 1 | Protect, prevent, prevention, trust, person, authorities, good, elderly, duty, world | 0.7376 | For the same reason as everyone else |
| 2 | Protect, family, virus, power, immunisation, efficacy, doctor, pandemic, others, president | 0.7409 | I trust doctors |
| 3 | Protect, desire, already, vaccine, protection, take, prevent, dose, prevention, world | 0.7540 | The vaccine affects everyone |

**Table 4:**
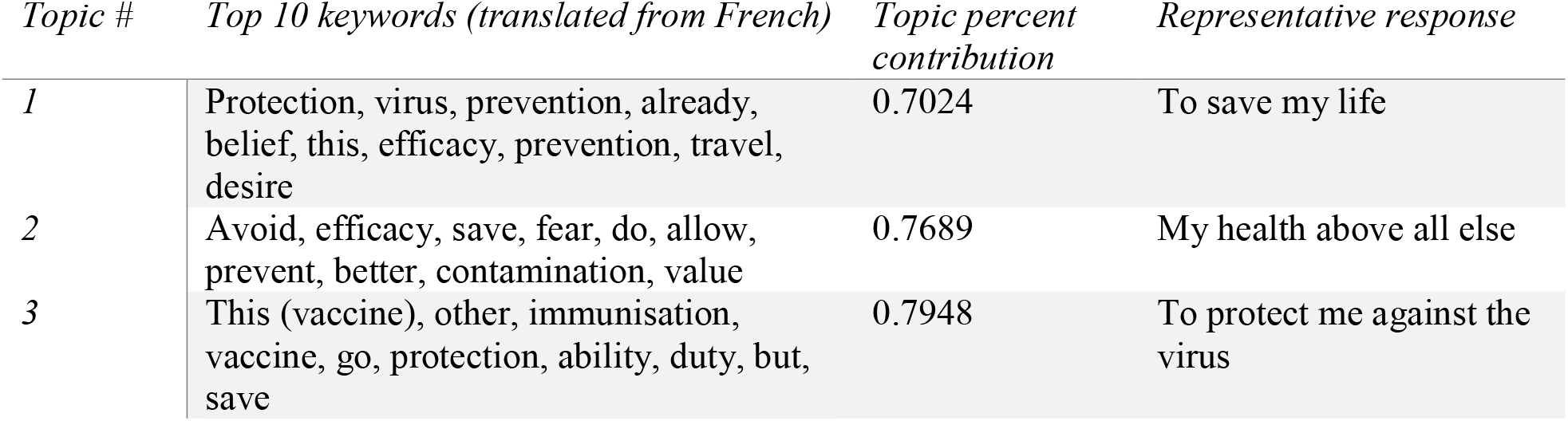
Reasons given for vaccine acceptance in Mali

## Conclusions

These findings point to high rates of vaccine hesitancy in Senegal and in Mali, particularly among youth, those with no chronic disease in the household, and those with less years of schooling. In both countries, the elderly appear significantly more likely to accept the vaccine, with only 42% in Senegal and 37% in Mali claiming to be hesitant.

There is significant overlap in terms when discussing reasons given both for vaccine hesitancy and vaccine acceptance in both countries, but particularly so for vaccine acceptance.

Respondents displayed three primary causes of hesitation - lack of belief in the importance of the vaccine, fear of the vaccine’s secondary effects, and general mistrust of the public health system. There did not appear to be significant differences in the causes for vaccine hesitancy between countries.

While the number of vaccine hesitancies over each dataset appear consistent (55% in Senegal and 52% in Mali), significantly more youth in Senegal appear hesitant than those in Mali (70% in Senegal and 57% in Mali). In Mali, women appear more hesitant (59%) than men (47%) to be vaccinated. This effect does not appear in Senegal (57% hesitancy for women and 60% for men).

While perceptions to vaccination may change over time, these results point to certain groups that may be targeted for public health communications campaigns. This research therefore hopes to inform national and international public health initiatives concerning both Senegal and Mali.

## Data Availability

All data for this research is available for academic purposes upon request.

## Footnotes

1 See : https://covid19.trackvaccines.org/country/senegal/

2 See : https://covid19.trackvaccines.org/country/mali/

## Notes

### Competing Interest Statement

The authors have declared no competing interest.

### Funding Statement

This research was funded by the International Development Research Centre (IDRC) in Canada.

### Author Declarations

Ethics for this research were approved by CNERS Senegal under the title: Protocole SEN20/91: "Usage de l'IA dans la lutter contre le COVID-19"

## Bibliography

Africa Centres for Disease Control and Prevention. https://africacdc.org/news-item/majority-of-africans-would-take-a-safe-and-effective-covid-19-vaccine/ (17 December 2020).

Blei D, Ng A, Jordan M. (2003). Latent Dirichlet Allocation. Journal of Machine Learning Research.

CORAF (2021). Les motifs de réticence vis-à-vis du vaccin anti-COVID-19 et les espaces de progression des options au Sénégal. White paper accessed at : http://www.coraf.org

MacDonald, N. E. (2015). Vaccine hesitancy: Definition, scope and determinants. Vaccine, 33(34), 4161–4164.

Machingaidze, S., Wiysonge, C.S. Understanding COVID-19 vaccine hesitancy. Nat Med 27, 1338–1339 (2021). https://doi.org/10.1038/s41591-021-01459-7

Meridith D, Sivry P. (2018). L’hésitation vaccinale et ses determinants. Exercer vol. 246.

Ratzan et al (2021). COVID-19 : A global survey shows worrying signs of vaccine hesitancy. The Conversation. https://theconversation.com/covid-19-a-global-survey-shows-worrying-signs-of-vaccine-hesitancy-148845

Reuters (2021). COVID-19 Tracking Dashboards for Senegal and Mali. Available at: https://graphics.reuters.com/world-coronavirus-tracker-and-maps/countries-and-territories/mali/

Ridde et al (2021). Au Sénégal, comment contrer la défiance envers la COVID-19. The Conversation. https://theconversation.com/au-senegal-comment-contrer-la-defiance-envers-le-vaccin-anti-covid-19-154863

Solís Arce, J.S., Warren, S.S., Meriggi, N.F. et al. COVID-19 vaccine acceptance and hesitancy in low-and middle-income countries. Nat Med 27, 1385–1394 (2021). https://doi.org/10.1038/s41591-021-01454-y

UNDP (2021). Vaccine equity dashboard. Website accessed at: https://data.undp.org/vaccine-equity/

Thiongane, O. (2021). En Afrique, la notion d’hésitation vaccinale est un modèle voyageur. The Conversation. https://theconversation.com/en-afrique-la-notion-dhesitation-vaccinale-est-un-modele-voyageur-158035

WHO (2021).Vaccine inequity is undermining the global economic recovery. News item accessed at: https://www.who.int/news/item/22-07-2021-vaccine-inequity-undermining-global-economic-recovery

